# The association between routine immunisation and COVID-19 vaccination in small island developing states

**DOI:** 10.1101/2024.12.28.24319741

**Authors:** Cyra Patel, Gizem Bilgin, Andrew Hayen, Martyn Kirk, Akeem Ali, Aditi Dey, Ginny Sargent, Meru Sheel

**Affiliations:** National Centre for Epidemiology and Population Health, Australian National University, Canberra ACT 2601, Australia; Sydney School of Public Health, Faculty of Medicine and Health, The University of Sydney, Camperdown NSW 2006, Australia; School of Public Health, University of Technology Sydney, Ultimo NSW 2007, Australia; World Health Organization, North Macedonia; National Centre for Immunisation Research and Surveillance, Sydney Children’s Hospitals Network, Westmead NSW 2145, Australia; Sydney Medical School, Faculty of Medicine and Health, The University of Sydney, Westmead NSW 2145, Australia; Sydney Institute for Infectious Diseases, Faculty of Medicine and Health, The University of Sydney, Westmead NSW 2145, Australia

**Keywords:** Immunisation, Vaccination, Small Islands Developing States, Health Systems, COVID-19 pandemic

## Abstract

**Objectives:** Understanding the link between routine immunisation (RI) performance and vaccination during an epidemic can provide insights on health systems resilience and investments to strengthen health systems. We examined the relationship between RI performance and COVID-19 vaccination coverage in small island developing states (SIDS).

**Methods:** COVID-19 vaccination coverage at four timepoints (June 2021, December 2021, June 2022 and December 2022) in 55 SIDS was our primary outcome. We examined associations with coverage of six childhood immunisations (5-year mean annual coverage for 2015–2019), pandemic-related disruptions to RI, new vaccine introductions, health system performance measures, and economic and demographic characteristics. We calculated Spearman correlations for continuous variables and mean COVID-19 vaccination coverage by categorical variables.

**Findings:** We found COVID-19 vaccination coverage was higher in countries that sustained RI coverage during the pandemic, and where HPV, influenza and measles-containing (second dose) vaccines had been introduced. There were weak correlations (|r|<0.4) between coverage of COVID-19 vaccination and RI, with a few exceptions of moderate correlations with the birth dose of hepatitis B vaccine (June 2022: r=0.421, p=0.007; December 2022: r=0.438, p=0.005) and first dose of measles vaccine (December 2021: r=0.420, p=0.002). COVID-19 vaccination coverage was strongly correlated with the density of physicians (June 2021: 0.897, p<0.001; December 2021: 0.785, p<0.001) and moderately correlated with that of nurses and midwives (June 2021: 0.630, p=0.001; December 2021: 0.605, p=0.002). COVID-19 vaccination coverage was lower in SIDS with lower country income and development status.

**Conclusions:** Countries that achieved high COVID-19 vaccination coverage maintained RI coverage, demonstrating health system resilience. Our findings highlight the importance of having sufficient skilled health professionals and experience in introducing new vaccines targeting different age groups into national programs, particularly in small island settings.

## Introduction

The Immunization Agenda 2030 (IA2030), the global vision and strategy for immunisation for the decade 2021 to 2030, emphasises the need to strengthen health systems to attain universal health coverage of all health services including immunisation.[1] Countries achieve and sustain high, equitable routine immunisation (RI) coverage when the various components of the immunisation systems align with fundamental health system components and work together effectively.[2,3] The recent advocacy and increasing investments in immunisation systems from disease-focused programs reflect this shift.[1,4] System-based approaches to achieving high, equitable RI coverage can have benefits like reaching underserved and vulnerable populations who previously had limited, if any, contact with the primary healthcare system.[5,6] However, there is limited evidence on the relationship between immunisation systems and the ability to deliver vaccines during public health emergencies caused by infectious disease outbreaks.

The COVID-19 pandemic provides an opportunity to apply a health systems lens to examine the link between RI performance and emergency vaccination. A recent analysis of COVID-19 vaccination coverage and immunisation program maturity found that COVID-19 vaccination coverage was 14% to 16% higher in countries with an adult seasonal influenza vaccination program.[7] However there was a high degree of heterogeneity in the health systems and political, socioeconomic and demographic contexts across countries, and in how COVID-19 vaccination programs were implemented.

In this study, we examined immunisation system factors associated with COVID-19 vaccination coverage within small island developing states (SIDS). SIDS are a group of 57 small, remote low-lying island nations, including 29 Caribbean nations, 19 Pacific Island Countries and Territories, eight Atlantic, Indian Ocean and South China Sea (AIS) member states, and one country in southeast Asia (Timor-Leste).[8] They are characterised by small population sizes and limited health infrastructure, especially highly constrained health workforces.[9–11] For example, in the Pacific region, Vanuatu, with a population of approximately 330,000, has 48 doctors and 353 nurses (0.16 and 1.16 per 1,000, respectively), while Niue has 3 doctors and 20 nurses (1.65 and 10.60 per 1,000, respectively) for their population of just under 2,000 people.[12] In the Caribbean, Dominica has 79 doctors and 461 nurses (1.12 and 6.51 per 1,000, respectively) for almost 73,000 people.[12] Their remoteness and economic and environmental vulnerability limit their capacity to respond rapidly to large infectious disease outbreaks. Many SIDS relied on border closures to control the importation of COVID-19.[13,14] Their small population sizes diminish their purchasing power with vaccine manufacturers, a significant disadvantage during the COVID-19 pandemic when supplies were limited and wealthy countries were buying up available stocks.[15,16] Health system organisation and performance vary across SIDS with different financing, political and socioeconomic influences. This was evident in responses to and outcomes of the COVID-19 pandemic, with some SIDS keeping the virus at bay until late 2021 and even 2022, while others experienced high case numbers and fatalities earlier on.[17–19] There is limited data on factors associated with COVID-19 outcomes, including vaccination coverage, in SIDS.

Examining immunisation system factors can provide insights into investments to immunisation systems and emergency vaccination. In this study, we examined the relationship between RI systems and COVID-19 vaccination coverage in SIDS.

## Methods

An immunisation system encompasses all components of a health system necessary to deliver vaccines, including all the organisations, institutions, resources, processes and activities involved in the delivery of immunisation programs.[20] In this study, we focused on the systems for routine immunisation and emergency vaccination, including the system’s capacity to deliver both concurrently, i.e. resilience. Key terms used in this study are defined in Box 1.

### Box 1

**Key definitions and assumptions**

- **Routine immunisation** comprises all vaccinations provided on a regular basis according to a country’s national vaccination schedule, including those delivered via mass vaccination campaigns (i.e. supplementary immunisation activities) to close gaps in coverage. Immunisation performance is most commonly measured by vaccination coverage, with higher coverage indicative of stronger performance.
- **Emergency vaccination** occurs to mitigate an infectious disease outbreak, using a novel vaccine antigen or a routine vaccine that targets different age and risk groups. This study focuses on COVID-19 vaccination during the acute phase of the

COVID-19 pandemic, i.e. between January 2020 and May 2023 when COVID-19 was considered a public health emergency of international concern.[53]

- **Health system resilience** is the capacity of the health system to prepare for and effectively respond to a crisis while maintaining core functions.[54] Resilient vaccination systems, therefore, are those that can provide emergency vaccination services in response to an infectious disease epidemic, with minimal disruptions to routine immunisation services.
- **New vaccine introduction** refers to the integration of newer and underutilised vaccines into a country’s national immunisation program. In this study, we focused on the second dose of measles containing vaccine (MCV2), pneumococcal conjugate vaccine (PCV), rotavirus vaccine and HPV vaccine as WHO recommends them for all countries, as well as influenza vaccine to examine the effect of integrating a vaccine targeting adulthood into national programs.

### Inclusion criteria

We included all SIDS (N=57) in line with the United Nations Office of the High Representative for the Least Developed Countries, Landlocked Countries and Small Island Developing States (UN-OHRLLS) classification including 39 WHO member states and 18 non-member states.[8] We included both WHO member and non-member states as they all experience social, infrastructural and economic challenges given their remote geography, small population sizes and vulnerability to climate change. We included all SIDS that had publicly reported data on COVID-19 vaccination coverage rates.

### Data sources

We extracted the following publicly available data (Table 1):

- Monthly COVID-19 vaccination coverage data for 2021 and 2022 from COVID-19 Vaccination Information Hub, collated from the WHO Joint Reporting Form (JRF) COVID-19 vaccination module.[21]
- WHO/UNICEF estimates of national immunisation coverage (WUENIC) data for the years 2015 to 2022, collated from countries annually via the JRF process. Data were sourced from the WHO Immunization Data Portal, including data on RI coverage, new vaccine introduction and immunisation system performance.[22] Data for RI coverage were not consistently reported for non-WHO member states, and were unavailable for new vaccine introduction status and immunisation system performance measures through the WUENIC system.
- World Bank Indicators data for health system performance (e.g. health worker density and childhood mortality rates) and country-level demographic characteristics (e.g. country population size).
- Country income level and development status from the United Nations Development Programme.

**Table 1:**
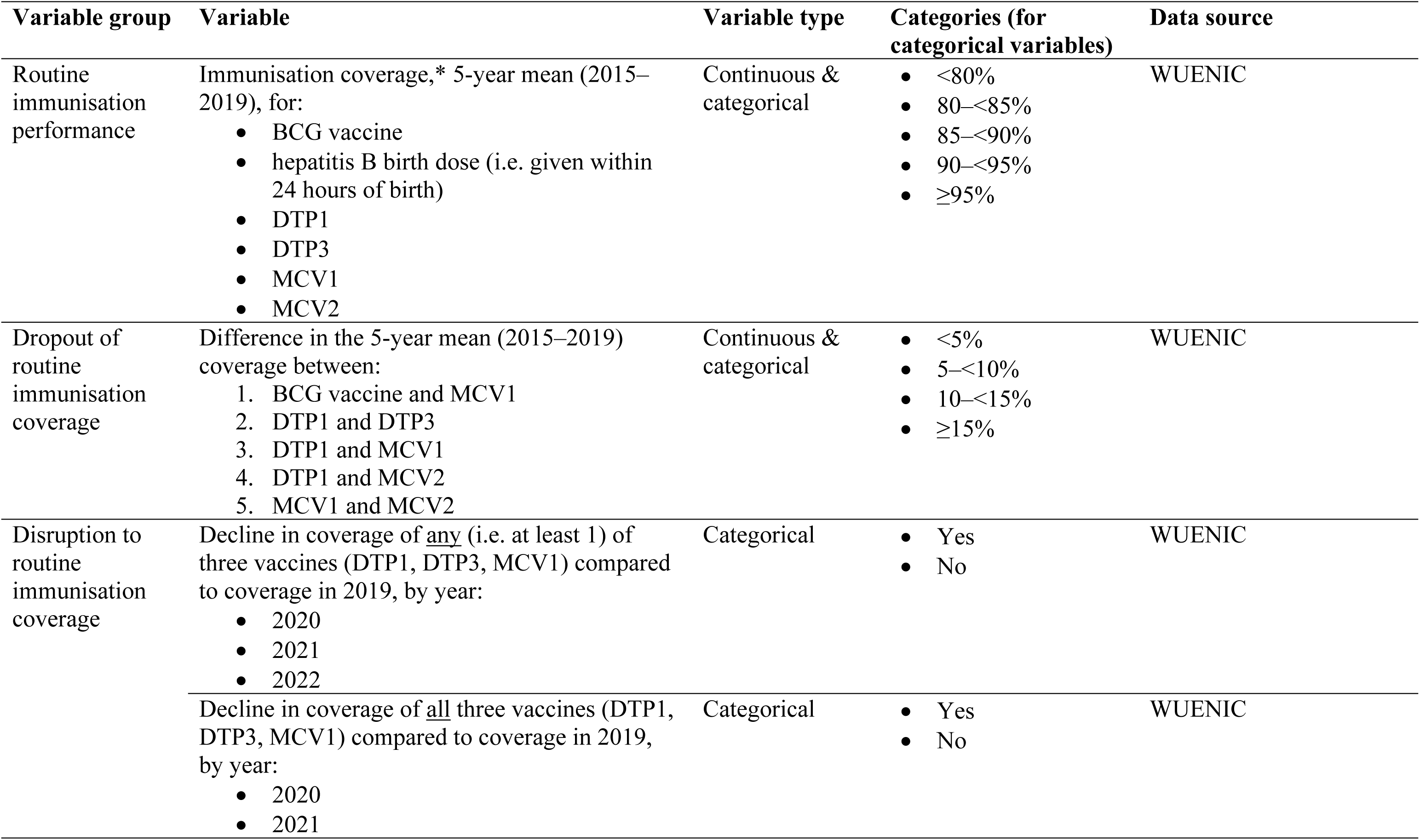
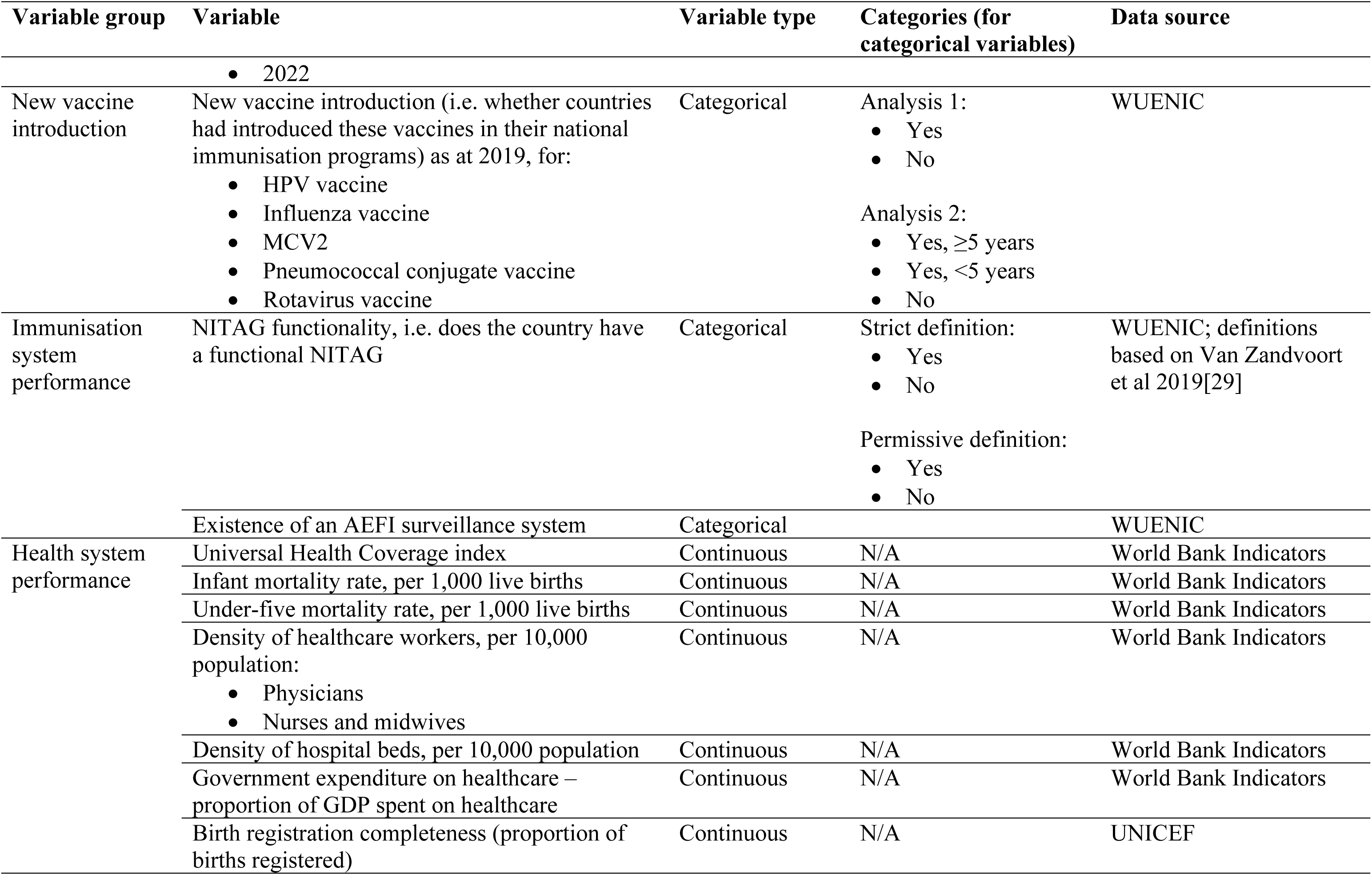
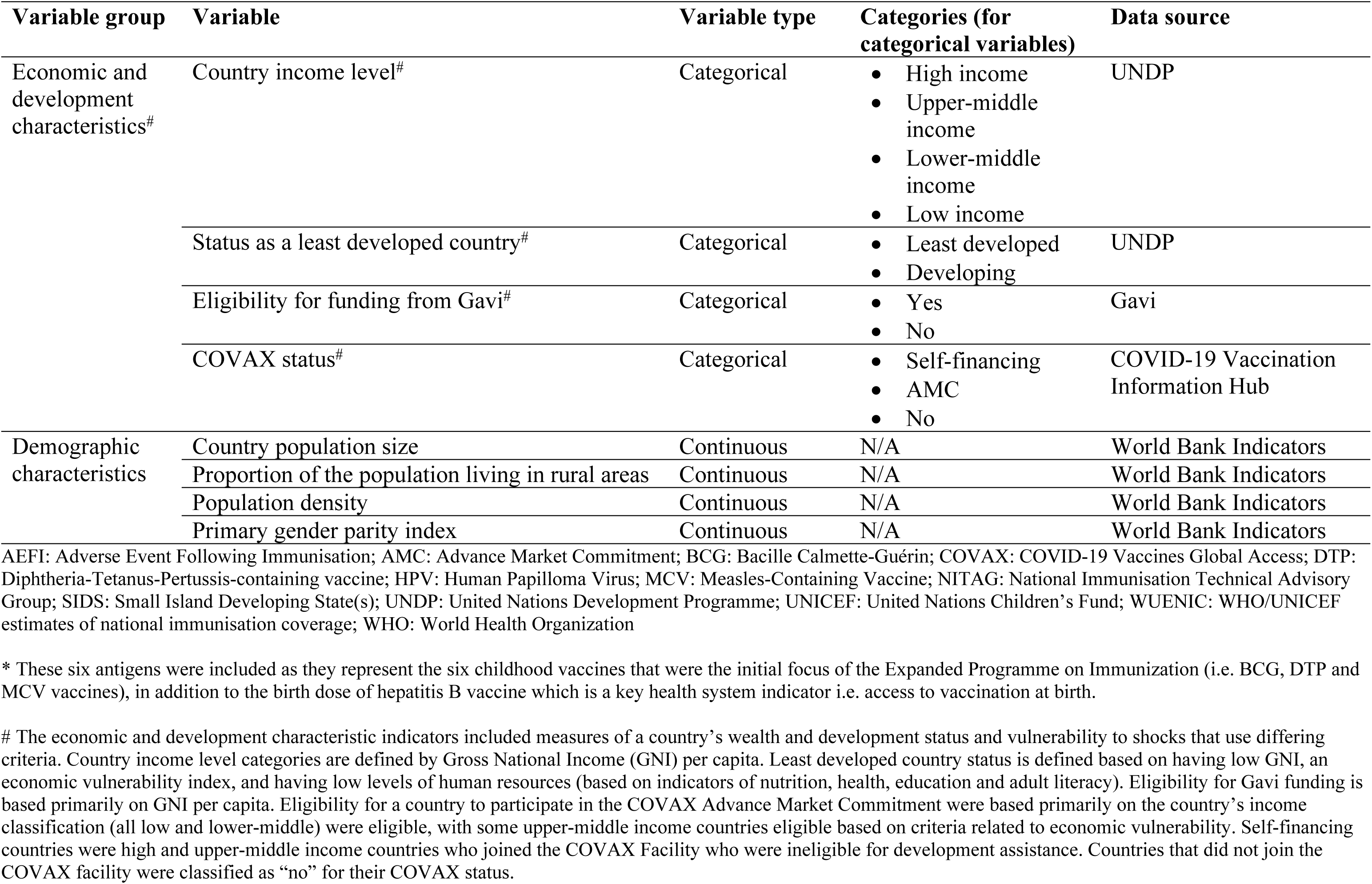
Independent variables included in the analysis and data sources.

All data were extracted in April 2024. As all data were aggregated at the country-level and obtained from publicly available sources, ethical approval was not required.

### Outcomes of interest

The primary outcomes of interest were coverage of: A) the first dose, and B) a complete primary course of COVID-19 vaccination coverage at four timepoints in the first two years of the global roll-out: 1) June 2021, 2) December 2021, 3) June 2022, and 4) December 2022. We did not include coverage for COVID-19 boosters in our study as we focused on vaccination efforts aimed at increasing population-level immunity during the acute phase of the pandemic.

### Independent variables: routine immunisation

Table 1 lists the definitions and details of independent variables. They included:

- The 5-year (2015–2019) mean annual coverage of six vaccines, i.e. Bacillus Calmette-Guérin (BCG) vaccine birth dose, hepatitis B vaccine birth dose, the first and third dose of diphtheria-tetanus-pertussis containing vaccine (DTP1 and DTP3), and the first and second dose of measles-containing vaccine (MCV1 and MCV2);
- Dropout of vaccination coverage, i.e. the proportion of children who did not receive subsequent doses of vaccination after receiving earlier doses in the series, which is often used as an indicator of the health system’s ability to consistently reach populations to achieve full vaccination;[3]
- Disruption to RI during the COVID-19 pandemic (i.e. 2020, 2021 and 2022), defined as decline in coverage of three vaccines (DTP1, DTP3 and MCV1) in comparison to 2019 (which has been used as a benchmark year in recent assessments of global immunisation coverage)[23–26] either alone or in combination;
- “New vaccine introduction” of five new and underutilised vaccines as of 2019, namely MCV2, PCV, rotavirus, HPV and influenza vaccines, as their introduction can reflect and impact health systems.[27]

### Independent variables: health system and macro factors

We included immunisation system performance measures that focus on system strengthening initiatives in recent years. These include the functionality of a national immunisation technical advisory group (NITAG)[28,29] and established national surveillance system for adverse events following immunisation (AEFI).[30,31] In line with our hypothesis that immunisation system performance is associated with health systems as a whole, we examined the relationship with universal health coverage (UHC) index, infant and under-five mortality rates, density of healthcare workers and hospital beds, government expenditure on healthcare and birth registration completeness.

We also included economic and development indicators (i.e. country income level, status as a least developed country, eligibility for funding from Gavi The Vaccine Alliance, and COVAX status), and demographic characteristics (i.e. country population size, the proportion of population living in rural areas, population density and the primary gender parity index) to account for macro-level factors.

Pre-pandemic data from 2019 was used for all health system and macro factors.

### Data analyses

We generated descriptive statistics using an exploratory data analysis approach to analyse the association between COVID-19 vaccination coverage and independent variables listed in Table 1. We calculated Spearman correlation where independent variables were continuous, and compared COVID-19 vaccination coverage by categorical variables. We generated Spearman correlations coefficients with p-values (p<0.05 considered significant) and 95% confidence intervals. We classified correlations as weak (|r| < 0.4), moderate (|r| ≥ 0.4 and |r| < 0.7) or strong (|r| ≥ 0.7).

We included all available data for SIDS that had COVID-19 vaccination coverage data, excluding those from specific analyses where data for specific independent variables were unavailable. In analyses with RI coverage, we used the mean of available coverage values (i.e. the denominator was the number of years for which data were available). We excluded SIDS from specific analyses only if they did not have any coverage values reported for a specific vaccine over the 5-year period. We limited the analyses by new vaccine introduction variables and immunisation system performance variables to WHO member states only (n=39), as data for non-member states were not available through WUENIC.

Analyses were conducted in RStudio (R version 4.4.1, 2024).[32]

## Results

Of the 57 SIDS, two (Martinique and the US Virgin Islands) did not have publicly-reported COVID-19 vaccination data and were excluded. Our final study dataset included 55 SIDS representing a mix of country income levels (see Table 2). The included SIDS were from the WHO regions of the Americas (n=27), Western Pacific (n=20), Africa (n=6) and Southeast Asia (n=2).

**Table 2:** Characteristics of included small island developing states (SIDS) (N=55)*

| **Characteristic** | **n** | **%** |
| --- | --- | --- |
| WHO region |  |  |
| Africa | 6 | 10.9% |
| Americas | 27 | 49.1% |
| South-East Asia | 2 | 3.6% |
| Western Pacific | 20 | 36.4% |
| SIDS region |  |  |
| AIS | 8 | 14.5% |
| Caribbean | 27 | 49.1% |
| PICs | 19 | 34.5% |
| N/A | 1 | 1.8% |
| Income level |  |  |
| Low | 1 | 1.8% |
| Lower-middle | 11 | 20.0% |
| Upper-middle | 16 | 29.1% |
| High | 22 | 40.0% |
| Other | 5 | 9.1% |
| Status as "Least developed country" |  |  |
| Least developed | 7 | 12.7% |
| Developing | 48 | 87.3% |
| Eligibility for Gavi funding |  |  |
| Yes | 7 | 12.7% |
| No | 32 | 58.2% |
| N/A (non-WHO member state) | 16 | 29.1% |
| COVAX membership |  |  |
| Self-financing | 12 | 21.8% |
| AMC | 23 | 41.8% |
| No | 4 | 7.3% |
| N/A (non-WHO member states) | 16 | 29.1% |

AIS: Atlantic, Indian Ocean and South China Sea; AMC: Advance Market Commitment; COVAX: COVID-19 Vaccines Global Access; PICs: Pacific Island Countries and Territories; SIDS: Small Island Developing State(s); WHO: World Health Organization
\* The 55 SIDS included. American Samoa, Anguilla, Antigua and Barbuda, Aruba, Bahamas, Barbados, Belize, Bermuda, British Virgin Islands, Cabo Verde, Cayman Islands, Commonwealth of Northern Marianas, Comoros, Cook Islands, Cuba, Curacao, Dominica, Dominican Republic, Fiji, French Polynesia, Grenada, Guadeloupe, Guam, Guinea-Bissau, Guyana, Haiti, Jamaica, Kiribati, Maldives, Marshall Islands, Mauritius, Micronesia (Federated States of), Montserrat, Nauru, New Caledonia, Niue, Palau, Papua New Guinea, Puerto Rico, Saint Lucia, Saint Vincent and the Grenadines, Saint Kitts and Nevis, Samoa, São Tomé and Príncipe, Seychelles, Singapore, Sint Maarten, Solomon Islands, Suriname, Timor-Leste, Tonga, Trinidad and Tobago, Tuvalu, and Vanuatu

### Associations with routine immunisation performance

We found weak correlations (r<0.4) between coverage of both the first dose and complete primary series of COVID-19 vaccination coverage and the 5-year mean annual coverage of RI included in the study, with few exceptions (Table 3, Appendix 1). These exceptions where there were moderate correlations (i.e. r≥0.4 but all were r<0.5) were for coverage of the birth dose of hepatitis B vaccine (r=0.425 [p=0.006, 95%CI: 0.056–0.730] and r=0.421 [p=0.007, 95%CI: 0.072–0.721] for the first dose and full primary series of COVID-19 vaccination coverage, respectively, in June 2022, and r=0.421 [p=0.007, 95%CI: 0.072–0.721] and r=0.438 [p=0.005, 95%CI: 0.098–0.715] for the first dose and full primary series of COVID-19 vaccination coverage, respectively, in December 2022) and the first dose of measles vaccine (r=0.420 [p=0.002, 95%CI: 0.145–0.664] for full primary series of COVID-19 vaccination and MCV1 in December 2021).

**Table 3:** Spearman correlations between COVID-19 vaccination coverage and 5-year (2015–2019) mean annual coverage of routine immunisations.

| Vaccine | N | June 2021 | December 2021 | June 2022 | December 2022 |
| --- | --- | --- | --- | --- | --- |
|  |  | rho | rho | rho | rho |
| Coverage of first dose of COVID-19 vaccination |  |  |  |  |  |
| BCG | 42 | 0.335* | 0.279 | 0.257 | 0.231 |
| DTP1 | 51 | 0.334* | 0.35* | 0.334* | 0.314* |
| DTP3 | 52 | 0.264 | 0.348* | 0.258 | 0.222 |
| HepB birth dose | 40 | 0.367* | 0.374* | **0.425\*\*** | **0.402\*** |
| MCV1 | 52 | 0.338* | 0.399** | 0.341* | 0.304* |
| MCV2 | 47 | 0.245 | 0.377** | 0.338* | 0.317* |
| Full coverage of primary series of COVID-19 vaccination |  |  |  |  |  |
| BCG | 42 | 0.309* | 0.294 | 0.242 | 0.230 |
| DTP1 | 51 | 0.311* | 0.392** | 0.360** | 0.345* |
| DTP3 | 52 | 0.255 | 0.378** | 0.311* | 0.250 |
| HepB birth dose | 40 | 0.279 | 0.369* | **0.421\*\*** | **0.438\*\*** |
| MCV1 | 52 | 0.267 | **0.420\*\*** | 0.373** | 0.334* |
| MCV2 | 47 | 0.178 | 0.393** | 0.359* | 0.349* |

\* p<0.05, \*\* p<0.01
Bolding shows show $|r|>0.4$

We observed moderate negative correlations (i.e. between –0.4 and –0.5) between COVID-19 vaccination coverage and dropout of coverage between routine vaccines, particularly between DTP1 and either MCV1 or MCV2, largely at the December 2021 timepoint (results in Appendix 2). The negative correlations indicate lower COVID-19 vaccination coverage with higher RI dropout. Correlations at all other timepoints were weak.

We found SIDS that experienced declines in RI coverage in 2021 and 2022 had lower COVID-19 vaccination coverage across the entire study period compared with those who did not experience disruptions (see Figure 1). This was true whether SIDS experienced declines for a single vaccine (DTP1, DTP3 or MCV1) or all three combined vaccines.

**Figure 1:**
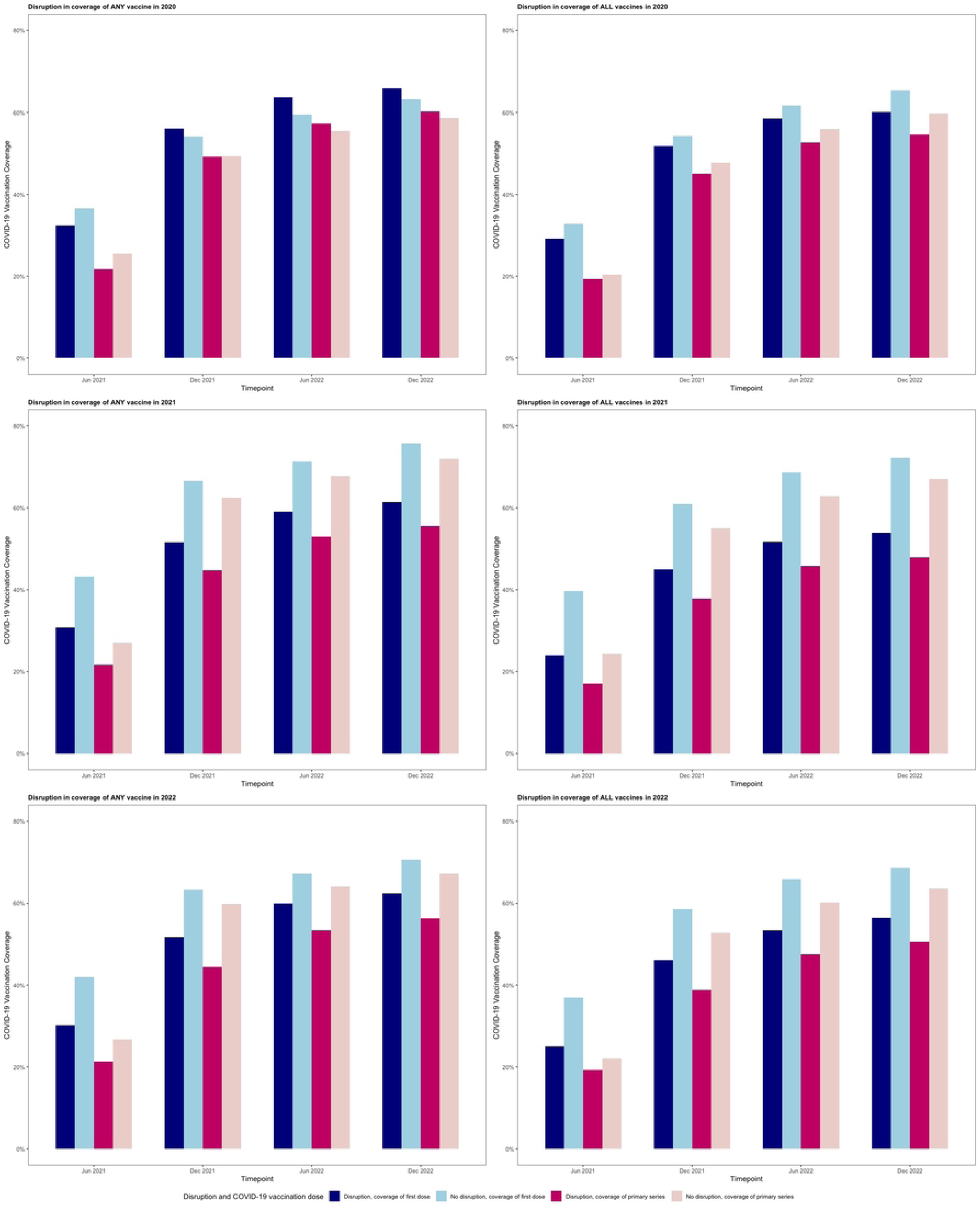
COVID-19 vaccination coverage by whether countries experienced declines in routine immunisation coverage (DTP1, DTP3 and MCV1) during the COVID-19 pandemic. The figures show COVID-19 vaccination coverage (first dose and primary series) at the four timepoints included in the study (i.e. June 2021, December 2021, June 2022 and December 2022), categorised by whether countries experienced disruptions to three routine immunisations (DTP1, DTP3 and MCV1) during the COVID-19 pandemic years (2020, 2021 and 2022). Notes for Figure 1:

- Disruption in ANY vaccine: declines in coverage of at least one of the three vaccines (DTP1, DTP3 or MCV1) were observed in the given year
- Disruption in ALL vaccines: declines in coverage of all three vaccines (DTP1, DTP3 or MCV1) were observed in the given year

### Association with new vaccine introductions

COVID-19 vaccination coverage was higher among SIDS that had introduced HPV, influenza and MCV2. These differences were greater in the earlier stages of the COVID-19 vaccination rollout, and reduced over time. When examined by years of introduction (Figure 2), SIDS that had introduced HPV vaccine and MCV2 vaccine within 5 years of 2019 had similar levels of COVID-19 vaccination coverage to those who had not introduced those vaccines as of 2019. COVID-19 vaccination coverage was higher for SIDS that had introduced HPV vaccine and MCV2 for 5 or more years. The reverse was observed for influenza vaccine introduction, with higher COVID-19 vaccination coverage among SIDS that had introduced influenza vaccine within 5 years, noting there were only three countries in this group (see Figure 2). No trends were observed regarding introduction of PCV or rotavirus vaccine (see Appendix 3).

**Figure 2:**
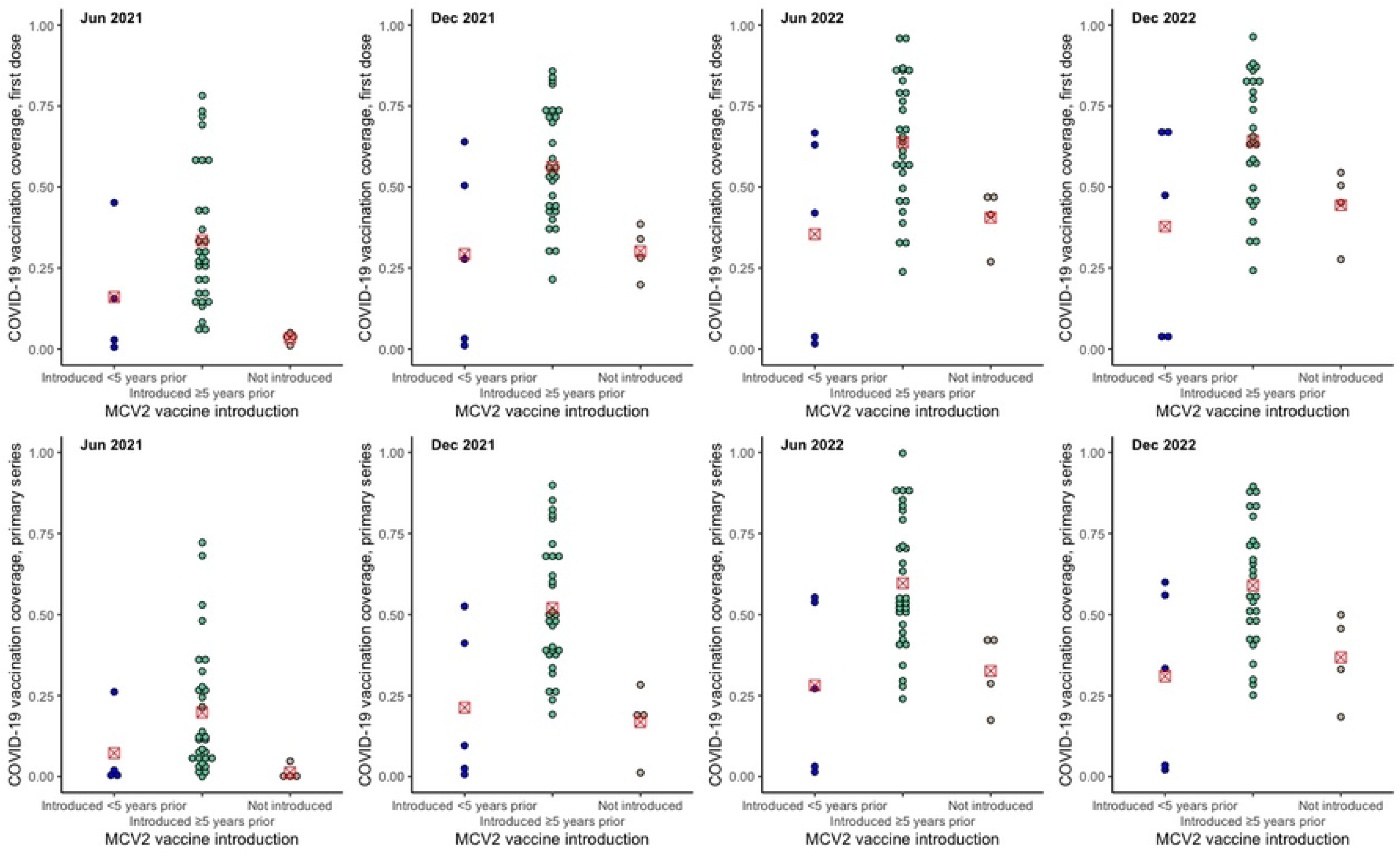
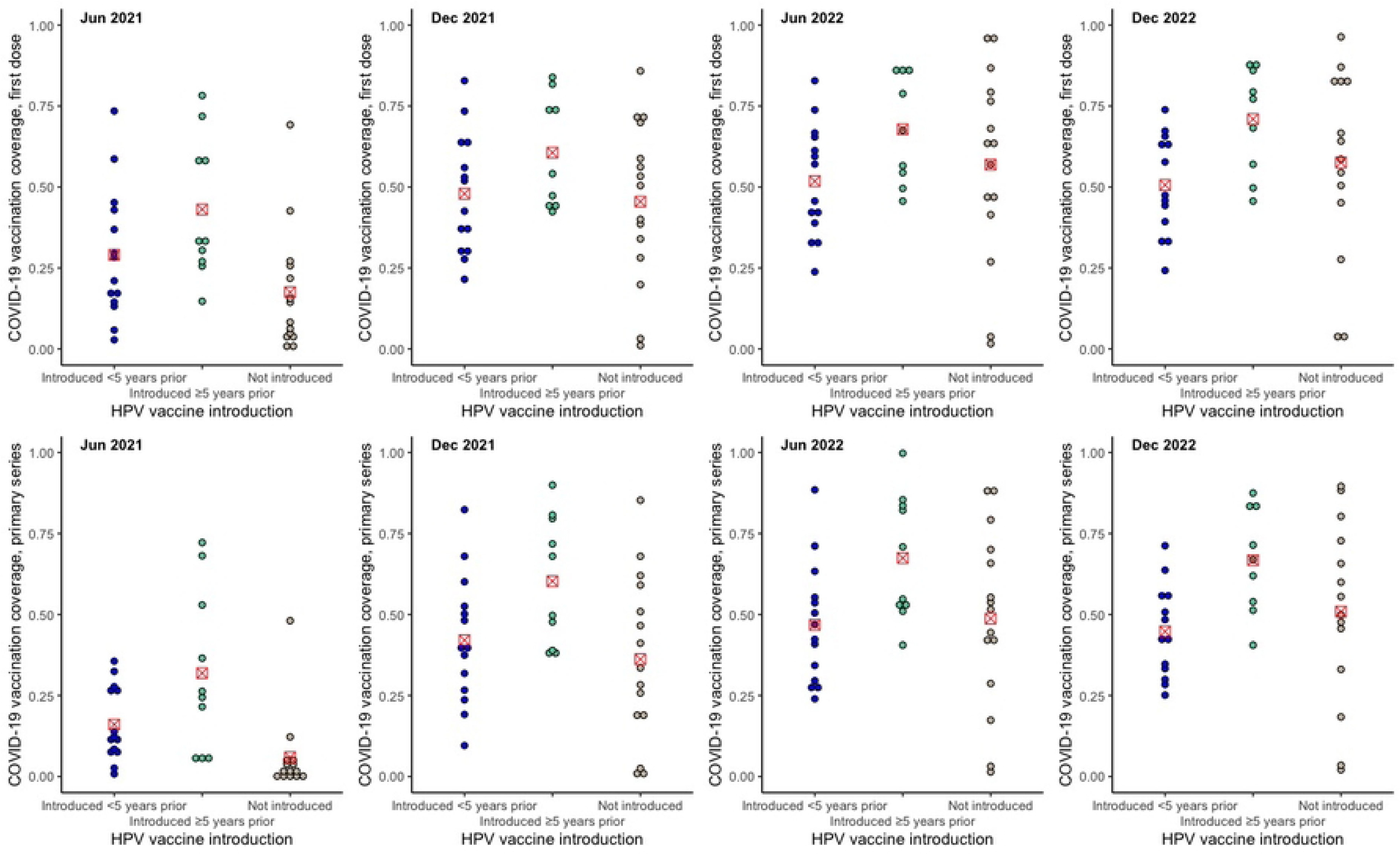
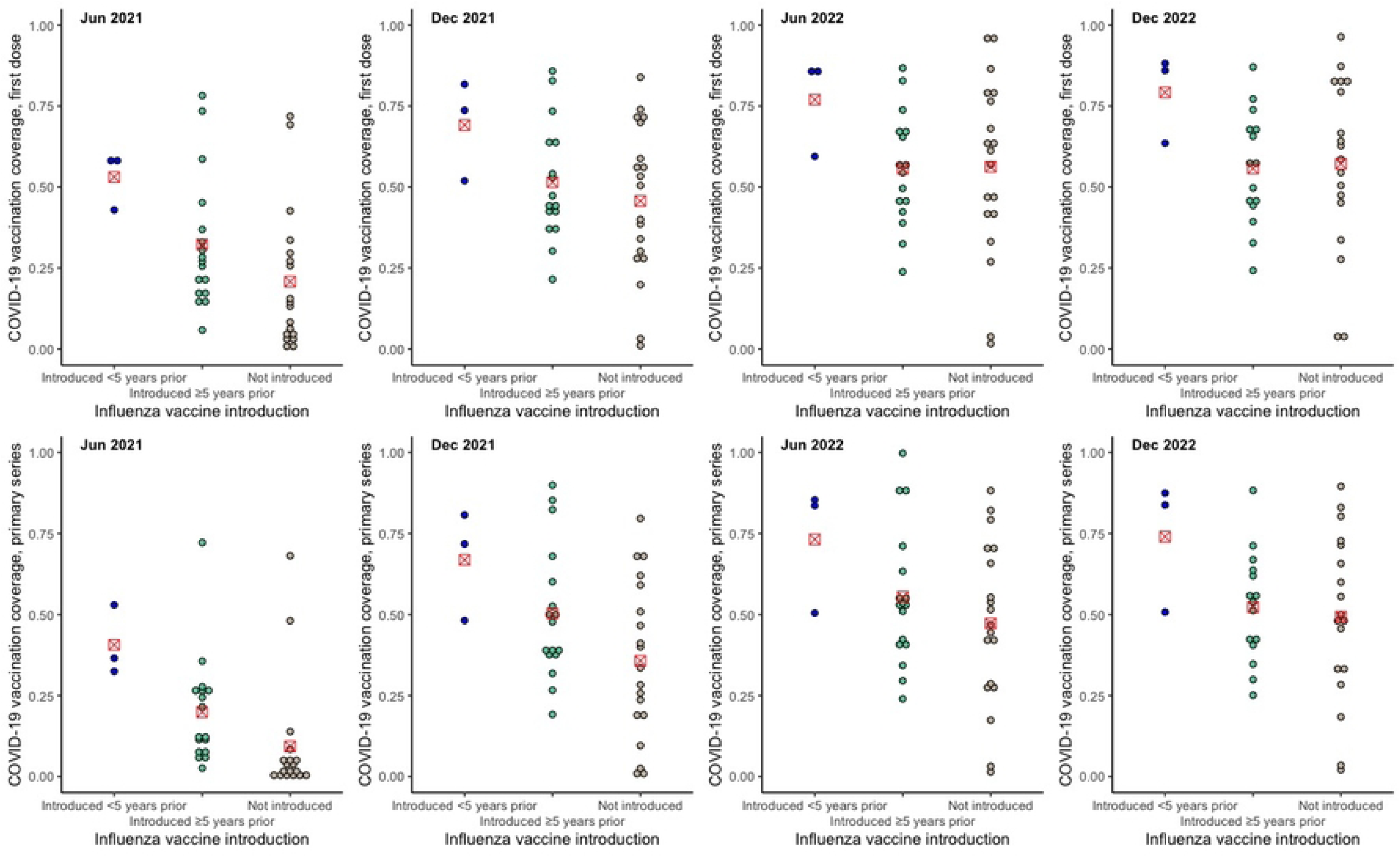
COVID-19 vaccination coverage by new vaccine introductions. Figures show COVID-19 vaccination by new vaccine introduction status: 1) introduced <5 years, 2) introduced ≥5 years, 3) not introduced. Dots represent COVID-19 vaccination coverage in each country, rounded to the nearest coverage point (e.g. 91.7% is rounded to 92%). The red marker represents mean COVID-19 vaccination coverage for the vaccine introduction category. 2A. MCV2 vaccine introduction 2B. HPV vaccine introduction 2C. Influenza vaccine

### Association with health system performance

We found a strong correlation between first dose and full COVID-19 vaccination coverage and the density of physicians, particularly in 2021 (first dose – June 2021: 0.905; December 2021: 0.759; primary series – June 2021: 0.897; December 2021: 0.785, p<0.001 for all), which declined over the study period to a moderate correlation in 2022 (first dose – June 2022: 0.643, p=0.001; December 2022: 0.608, p=0.002; primary series – June 2022: 0.654, p=0.001; December 2022: 0.625, p=0.001 (see Table 4 and Appendix 4; Appendix 5 for graphs). We observed a moderate correlation between first dose and full COVID-19 vaccination coverage and the density of nurses and midwives, which was more closely correlated in 2021 than in 2022 (see Table 4).

**Table 4:** Spearman correlations between COVID-19 vaccination coverage and density of health resources (workforce and hospital beds)

| Vaccine | June 2021 | December 2021 | June 2022 | December 2022 |
| --- | --- | --- | --- | --- |
|  | rho | rho | rho | rho |
| **Coverage of first dose of COVID-19 vaccination** |  |  |  |  |
| Physicians per 1,000<sup>#</sup> | **0.905\*\*\*** | **0.759\*\*\*** | **0.643\*\*** | **0.608\*\*** |
| Nurses and midwives per 1,000 | **0.598\*\*** | **0.517\*** | 0.389 | 0.360 |
| Hospital beds per 10,000 | **0.581\*\*\*** | 0.312 | 0.290 | 0.297 |
| **Full coverage of primary series of COVID-19 vaccination** |  |  |  |  |
| Physicians per 1,000* | **0.897\*\*\*** | **0.785\*\*\*** | **0.654\*\*** | **0.625\*\*** |
| Nurses and midwives per 1,000 | **0.630\*\*** | **0.605\*\*** | 0.429* | 0.430* |
| Hospital beds per 10,000 | **0.641\*\*\*** | 0.370* | 0.314 | 0.315 |

\* p<0.05, \*\* p<0.01, \*\*\* p<0.001
Bolding shows show $|r|>0.4$
Shading and folding shows strong correlations i.e. $|r|>0.7$
<sup>#</sup> One country was an outlier and excluded from this analysis. Correlation values were strong in a sensitivity analysis that included the outlier country.

We did not observe any clear associations between COVID-19 vaccination coverage and other health systems variables included in this study, including having a functional NITAG, the proportion of births registered, UHC index, government expenditure on healthcare, and under-5 and infant mortality rates (see Appendices 6 and 7). While some statistically significant moderate correlations were observed, visual inspection of scatter plots showed clustering of datapoints with correlations driven by a handful of datapoints (see Appendix 7).

### Association with country-level characteristics (economic, development and demographic factors)

COVID-19 vaccination coverage was higher in SIDS with higher country income during the period from January 2021 to December 2022. Coverage was highest in high-income countries, followed by upper-middle, lower-middle and low-income countries (see Figure 3, and Table 5). COVID-19 vaccination coverage was lower among SIDS classified as a “least developed country” during the same period, and in those eligible to receive Gavi funding (Table 5). COVID-19 vaccination coverage was highest in the four SIDS who did not participate in the COVAX facility, followed by self-financing members and AMC members (Table 5).

**Figure 3:**
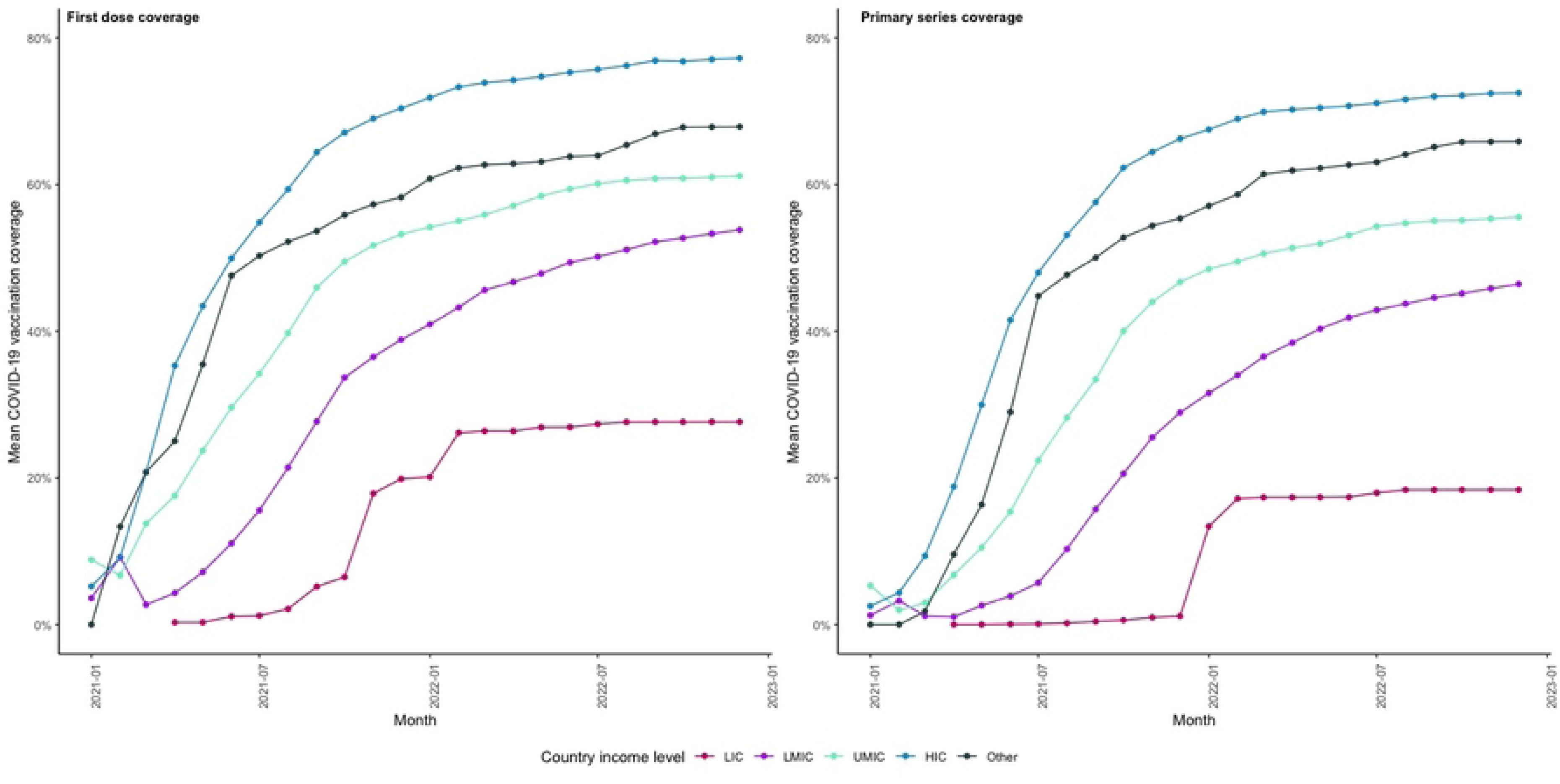
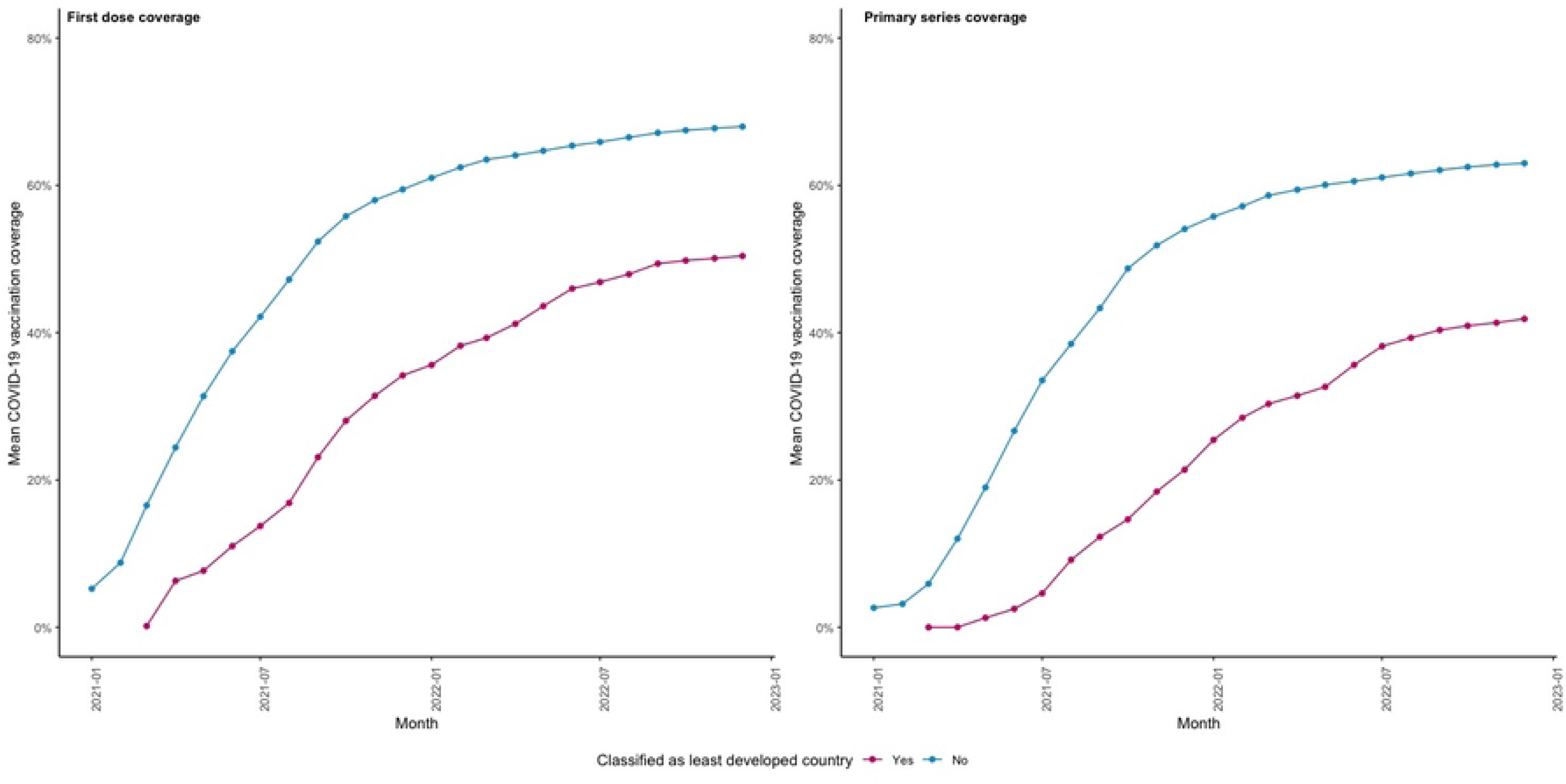
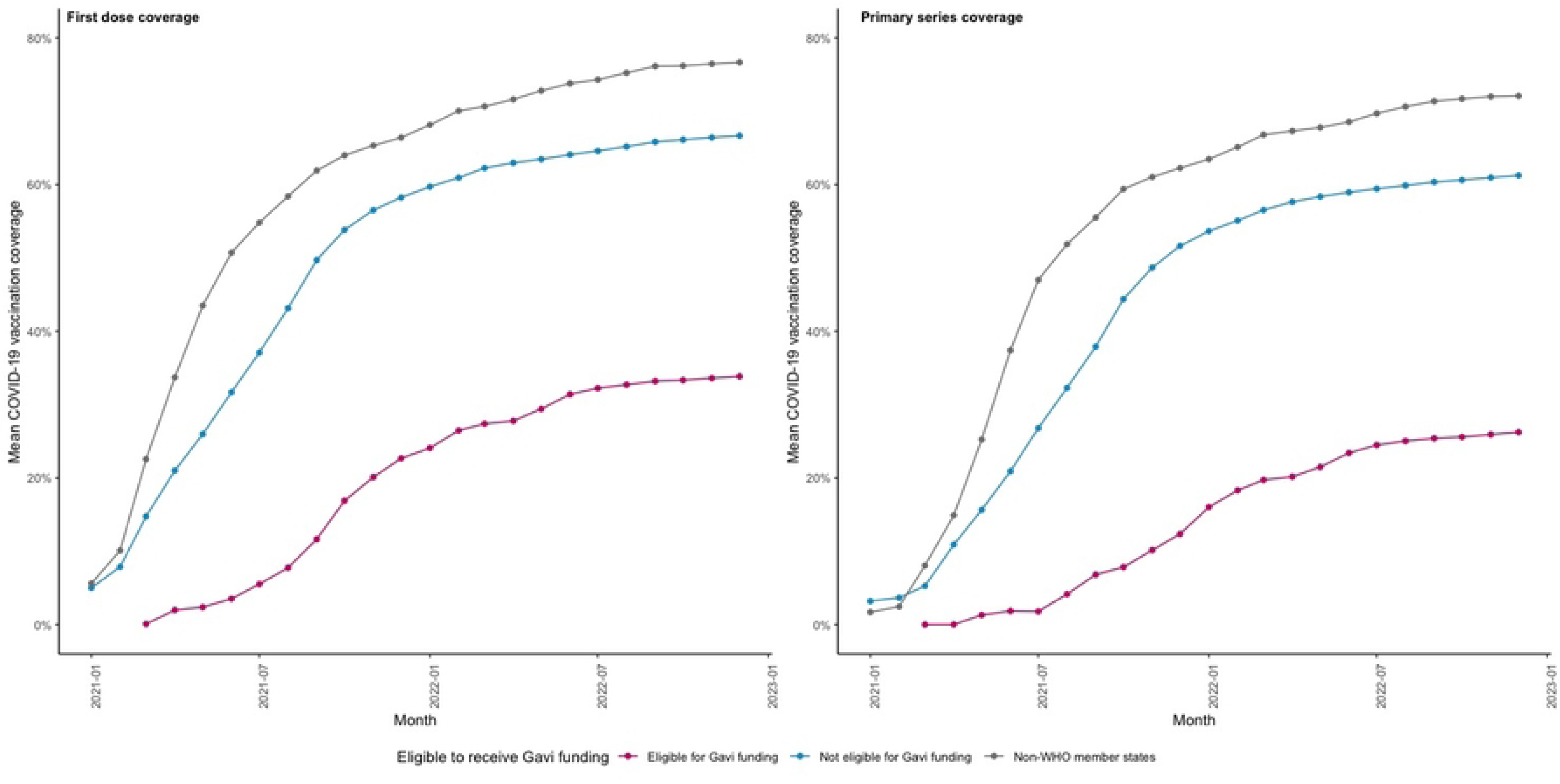
Monthly COVID-19 vaccination coverage by economic factors. Figures show monthly COVID-19 vaccination coverage (first dose and primary series) by various economic factors, namely country income level, status as a least developed country, and eligibility for Gavi funding. 3A. Income level 3B. Least developed status 3C. Eligibility for Gavi funding

**Table 5:** COVID-19 vaccination coverage at four timepoints in 2021 and 2022, by economic factors.

| Characteristic | N | Coverage of first dose of COVID-19 vaccination |  |  |  | Coverage of primary series of COVID-19 vaccination |  |  |  |
| --- | --- | --- | --- | --- | --- | --- | --- | --- | --- |
|  |  | Jun 2021 | Dec 2021 | Jun 2022 | Dec 2022 | Jun 2021 | Dec 2021 | Jun 2022 | Dec 2022 |
| Income level |  |  |  |  |  |  |  |  |  |
| Low | 1 | 1.1% | 19.9% | 26.9% | 27.6% | 0.1% | 1.2% | 17.4% | 18.4% |
| Lower-middle | 11 | 10.8% | 38.5% | 49.4% | 54.2% | 3.9% | 27.9% | 42.0% | 46.8% |
| Upper-middle | 16 | 29.4% | 53.4% | 60.0% | 61.6% | 16.2% | 47.5% | 53.5% | 56.1% |
| High | 22 | 47.6% | 68.3% | 73.2% | 75.7% | 38.7% | 63.6% | 68.5% | 70.6% |
| Other | 5 | 47.6% | 58.3% | 63.8% | 67.9% | 29.0% | 55.4% | 62.7% | 65.9% |
| LDC status |  |  |  |  |  |  |  |  |  |
| Yes | 7 | 11.0% | 34.2% | 46.0% | 50.4% | 2.5% | 21.4% | 35.6% | 41.9% |
| No | 48 | 37.5% | 59.4% | 65.4% | 68.0% | 26.7% | 54.1% | 60.6% | 62.9% |
| Eligibility for Gavi funding |  |  |  |  |  |  |  |  |  |
| Yes | 7 | 3.5% | 22.7% | 31.4% | 33.8% | 1.9% | 12.4% | 23.4% | 26.2% |
| No | 31 | 31.7% | 58.2% | 64.1% | 66.7% | 20.9% | 51.6% | 58.9% | 61.2% |
| N/A* | 17 | 50.7% | 66.4% | 73.8% | 77.2% | 37.4% | 62.3% | 68.5% | 72.4% |
| COVAX status |  |  |  |  |  |  |  |  |  |
| AMC | 23 | 18.7% | 42.7% | 52.1% | 55.6% | 8.4% | 33.6% | 44.4% | 48.4% |
| Self-financing | 12 | 37.2% | 57.8% | 63.4% | 65.9% | 24.8% | 53.5% | 58.1% | 61.0% |
| No | 4 | 56.3% | 81.6% | 85.4% | 90.4% | 36.1% | 79.8% | 85.6% | 89.1% |
| N/A | 16 | 48.9% | 68.2% | 72.5% | 74.0% | 41.8% | 63.2% | 68.5% | 69.5% |

LDC = Least Developed Country
\* Non-member WHO states

No clear associations were observed between demographic factors including proportion of population in rural areas, population density and population size, or the primary gender parity index (see Appendix 8).

## Discussion

Our analysis of immunisation in SIDS provides critical insights into the relationship between RI and vaccination during infectious disease epidemics. We found that sustaining pre-pandemic RI coverage rates during the pandemic was positively associated with having high COVID-19 vaccination coverage. Positive associations were also found between COVID-19 vaccination coverage and other immunisation and health system performance factors particularly health workforce density, having introduced newer vaccines, and economic and development factors. In contrast, there were mostly weak associations between COVID-19 vaccination coverage and RI coverage rates achieved prior to the COVID-19 pandemic in SIDS.

The weak associations between COVID-19 vaccination and level of RI coverage prior to the pandemic is not entirely surprising, since the speed, scale and target population of COVID-19 vaccination programs differed to those of RI programs which usually focusses on children under 5 years of age. It is consistent with results of earlier assessments which reported an absence of a relationship between national immunisation capacity and readiness to implement COVID-19 vaccination programs[33], and between the maturity of childhood immunisation programs and COVID-19 vaccination coverage.[7] The moderate associations that we observed between COVID-19 vaccination and RI vaccination coverage were largely with coverage of the birth dose of hepatitis B vaccine. This is typically given in a health facility post-delivery, which differs from vaccines given at other timepoints in infancy which are delivered through a combination of fixed, outreach and mobile vaccination services, implying access to health facilities may have been an important factor for attaining COVID-19 vaccination coverage in SIDS. We also observed moderate negative correlations between COVID-19 vaccination coverage and dropout of annual coverage for RI in December 2021, when COVID-19 vaccination supply constraints were easing. This may indicate the role of disparities in wealth, education and other socioeconomic factors, which are predictors of vaccination dropout, in achieving high and equitable coverage during an epidemic.[34]

The finding that SIDS that did not experience declines in RI coverage during the pandemic achieved high COVID-19 vaccination coverage aligns with the principles of a “strong” health system being a resilient one. Resilience is dependent on having a high functioning and adaptive health system, underpinned by equitable access to essential health services.[35] However, we did not find an association between universal health coverage and COVID-19 vaccination coverage, contrary to a recent study that demonstrated countries with higher universal health coverage index scores had higher rates of COVID-19 vaccination.[36] This might reflect intensified international development partner efforts in response to COVID-19 especially in lower income countries, or the public response to COVID-19 control measures, such as requirements for vaccination certificates for travel.[37]

The association between COVID-19 vaccination coverage and the density of medical doctors was the strongest in our analysis, with a weaker association with the density of nurses and midwives observed. This highlights the importance of having sufficient skilled health professionals to meet the surge requirements of a health emergency. Limited workforce quantity and capability were key barriers in COVID-19 vaccination program implementation globally.[38,39] Many countries had to diversify their workforce by recruiting students and retired health professionals.[40] Modelling studies found an additional 744,000 health workers were needed globally, largely in low-income countries, to achieve 70% coverage for COVID-19 vaccination in all countries by mid-2022 at a cost of US$2.5 billion.[39] While workforce capacity is critical for health system resilience in all settings, it is possibly more important in SIDS where the small numbers of health professionals greatly limits surge capacity,[10,12,41] and where workers take on multiple roles even under normal circumstances.[42] In these settings, a small number of additional health workers could have impacted countries’ ability to deliver immunisation services.

Similar to other studies,[43–46] we found a clear association between COVID-19 vaccination coverage and country income level. Least developed countries, with weaker health systems,[47] had lower COVID-19 vaccination coverage throughout the study period. It also reflects procurement and access to COVID-19 vaccines with low-and-middle-income countries unable to out-compete wealthy countries in negotiations with manufacturers.[44] For SIDS, vaccine logistics are more challenging and expensive due to geographically remote and dispersed islands. Small population sizes in SIDS also diminishes their purchasing power.[11,48] We found inequities in coverage persisted throughout 2022 when COVID-19 vaccine supply was no longer an issue, indicating health systems factors other than purchasing power and access to vaccine supplies contribute and are critical for vaccine delivery during epidemics. This includes workforce density, which is associated with country income.[39]

The relationship with introduction of new vaccines (e.g. influenza, HPV and MCV2) aligns with previously published data that showed an association between COVID-19 vaccination coverage and having an adult seasonal influenza program.[7] New vaccine introduction requires all components of the immunisation system to work together, including policy decision financing and vaccine procurement, cold chain capacity, workforce training and community engagement.[49,50] Furthermore, the associations we found were with vaccines introduced to age groups beyond the first year of life. This indicates that system strengthening occurs through the introduction of new systems and processes needed to deliver vaccines to a target population other than infants, and especially when the target population is in adolescence or adulthood. Similarly, the relationship with years since introduction suggests that it takes time for countries to adjust and embed new vaccine delivery processes into health systems.

Our analysis was limited by the lack of publicly-available coverage data for all routinely-administered vaccines in SIDS, particularly for non-WHO member states. Due to the small number of SIDS in some categories, we were unable to conduct regression analyses and control for potential confounders such as country income level. Nonetheless, our descriptive study reveals important information on the link between emergency vaccination and health and immunisation systems in SIDS.

While our study focussed on disruptions to RI, future research to identify and measure health system factors predictive of resilient immunisation systems would be useful. Additional factors that could be examined include service delivery and accessibility, vaccine acceptance and demand, and cold chain capacity, which were beyond the scope of this study. Cultural and political factors also have a role, with multiple studies reporting that trust in government was strongly associated with higher, faster and earlier uptake of COVID-19 vaccination and other outcomes during the pandemic including COVID-19 mortality.[7,51,52] Other factors, such as policies related to mandatory vaccination requirements for work or travel, could have also influenced COVID-19 vaccine uptake. Qualitative methods could be used to explore other health systems characteristics such as flexibility and adaptability, which are difficult to quantify.

## Conclusions

Our study provides support that high-performing health systems are resilient in achieving public health outcomes like high vaccination coverage during an epidemic while sustaining routine vaccination. They are underpinned by a health workforce, sufficient in both quantity and capability, and a flexible system able to adapt existing service delivery models quickly to meet emergency needs. Our study provides insights on where system strengthening efforts can focus to prevent shocks to health services not just for immunisation but primary health care in general.

## Acknowledgements

We would like to acknowledge the statistical training services provided by Biological Data Science Institute at the Australian National University and Sydney Informatics Hub at the University of Sydney.

## Authors’ contributions

CP, MS and GS conceptualised the study. CP designed the analysis plan with agreement from all authors. CP and GB conducted the analyses. CP drafted and revised the manuscript with critical input from all authors. All authors read and approved the final manuscript.

## Ethical approval and consent to participate

No ethical approval was required as this study used publicly available datasets on national-level data.

## Data availability

Data were obtained from publicly available datasets. All analysed findings have been presented in the manuscript and supplementary files. Corresponding authors may be contacted for additional information.

## Supporting information

S1: Spearman correlations between COVID-19 vaccination coverage and 5-year (2015–2019) mean annual coverage of routine immunisations

S2: Spearman correlations between COVID-19 vaccination coverage and dropout of annual coverage (5-year mean, 2015–2019) of routine immunisations

S3: COVID-19 vaccination coverage by new vaccine introductions

S4: Spearman correlations between COVID-19 vaccination coverage and density of health resources (workforce and hospital beds)

S5: Scatterplots of COVID-19 vaccination coverage and workforce density

S6: COVID-19 vaccination coverage at four timepoints in 2021 and 2022, by health system variables included in the study

S7: Scatterplots of COVID-19 vaccination coverage and health system variables included in the study

S8: Scatterplots of COVID-19 vaccination coverage and demographic factors included in the study

